# Time To Navigate (TTN): A practical objective clinical measure for freezing of gait severity in people with Parkinson’s disease

**DOI:** 10.1101/2023.08.18.23294294

**Authors:** A.E. Scully, D. Tan, B.I.R. de Oliveira, K.D. Hill, R. Clark, Y.H. Pua

**Author notes:** Address all correspondence to Dr Scully at:.

## Abstract

**Objectives:** Existing objective assessments for freezing of gait (FOG) severity may be unwieldy for routine clinical practice. To provide an easy-to-use clinical measure, this cross-sectional study explored if time to complete the recently-validated FOG Severity Tool (or its components) could be used to reflect FOG severity.

**Methods:** People with Parkinson’s disease who could independently ambulate eight-metres, understand instructions, and without co-morbidities severely affecting gait were consecutively recruited from outpatient clinics. Participants were assessed with the FOG Severity Tool in a test-retest design, with time taken for each component recorded using a stopwatch during video-analysis. Validity of total FOG Severity Tool time, time taken to complete its turning and narrow-space components (i.e., Time To Navigate, TTN), and an adjusted-TTN were examined through correlations with the FOG Questionnaire, percentage of time spent with FOG, and FOG Severity Tool-Revised score. To facilitate clinical interpretation, TTN cutoff was determined using scatterplot smoothing (LOESS) regression whilst minimal important change (MIC) was calculated using predictive modelling.

**Results:** Thirty-five participants were included [82.9%(n=29)male; Median(IQR): age – 73.0(11.0)years; disease duration – 4.0(4.5)years]. The FOG Severity Tool time, TTN, and adjusted-TTN similarly demonstrated moderate correlations with the FOG Questionnaire and percentage-FOG, and very-high correlations with FOG Severity Tool-Revised. TTN was nonlinearly related to FOG severity such that a positive relationship was observed in the first 300-seconds, beyond which the association plateaued. MIC for TTN was 15.4-seconds reduction in timing (95%CI 3.2 to 28.7).

**Conclusions:** The TTN is a feasible, interpretable, and valid test of FOG severity, demonstrating strong convergent validity with the FOG Severity Tool-Revised. In busy clinical settings, TTN provides a viable alternative when use of existing objective FOG measures is (often) unfeasible.

**Impact statement:** Busy clinicians need easy-to-use measures. In under 300-seconds, TTN test offers this for FOG severity, with a 15.4-seconds decrease in TTN time considered minimal improvement.

## Introduction

Freezing of gait (FOG) is defined as an episodic inability to effectively progress the feet despite an intention to advance.1 It is a disabling motor impairment, commonly seen in people with Parkinson’s disease.2 People with Parkinson’s disease and FOG have 3.5 times the odds of functional dependence compared to those with Parkinson’s disease without FOG.3 Regardless of disease severity, self-reported FOG severity has demonstrated significant, moderately-strong associations with self-reported disability.4,5 FOG is also a major cause of falls in people with Parkinson’s disease.6,7

Adequate assessments for FOG severity are crucial in the clinical context for guiding treatment decisions, evaluating treatment effectiveness, and monitoring disease progression.^8,9^ Self-reported outcomes for FOG severity have limitations, so current recommendations are to include objective evaluations too.^8,9^ In research, the percentage of time spent with FOG during performance of FOG-triggering tasks is considered the “gold standard” outcome.^10^ This requires video annotations,^10^ which is unfeasible for routine clinical practice. From as early as 1993, clinician-rated FOG severity outcomes have been published – some without validity and reliability investigated,^11,12^ some with only reliability,^13–15^ and some with both validity and reliability reported.^16–19^ However, none have translated to routine clinical practice, with clinicians expressing reasons of perceived impracticality and lack of usefulness.^9,20^

Recently, we developed a clinician-rated FOG severity outcome through a Delphi study,^20^ validated it,^21^ and improved on it to allow better differentiation of severity of FOG types beyond trembling-in-place and complete akinesia.^22^ The median completion time was 6.5 minutes (range: 3.5 to 16.0).^21^ For the recently-validated FOG Severity Tool–Revised scoring method, severity of shuffling and festination (based on number of steps taken) and trembling-in-place and complete akinesia (based on frequency and duration; i.e., whether there were one or multiple very brief, short, medium, or long episodes) were each rated on five-point scales and summed.^22^ Apart from measurement properties like validity and reliability, according to the COnsensus-based Standards for the selection of health Measurement INstruments (COSMIN), feasibility and interpretability are equally important factors for outcome measure selection.^23,24^ Feasibility is the ease of application, including completion time, costs, and type and ease of administration.^23^ Interpretability is the extent to which a qualitative meaning can be assigned to the quantitative scores, with the COSMIN recommending minimal important change (MIC) for interpretability of change in scores.^23^ To facilitate implementation in routine clinical practice, we explored scoring methods beyond clinician-rated scales in consideration of simplifying the outcome for better feasibility.

Aside from the percentage of time spent with FOG,^25–27^ previous studies have directly quantified FOG severity with number of FOG episodes,^25–30^ number of steps,^31^ and time taken to complete the FOG-triggering tasks.^28,31–33^ Several studies used methods less feasible for routine clinical practice [i.e., video annotations,^25–27,29^ inertial measurement units^31^] and some did not investigate validity.^26,28–31^ Number of FOG episodes has been shown to have poorer reliability due to varied definitions of when a single FOG episode starts and ends.^25,26^ For step count, we demonstrated its use to quantify severity of shuffling and festination in the FOG Severity Tool– Revised.^22^ Though validity was shown through its sufficient correlation with self-reported FOG severity,^22^ recording the number of steps taken in a clinical setting requires additional attention throughout the task. Moreover, counting steps can be more challenging during instances of festination or loss of gait rhythmicity. Conversely, using time taken can ease implementation as less expertise and training is needed.^32,34^

The time taken to complete Ziegler’s FOG score,^19^ as measured with a stopwatch, has been suggested as a proxy measure of FOG severity,^32,33^ though its interpretability is unclear. With videos of ten people with Parkinson’s disease and moderate-to-severe FOG, Goh and colleagues demonstrated moderate correlation between time taken and total percentage of time spent with FOG (r = 0.67).^33^ This was not unexpected since percentage of time spent with FOG is derived from time taken (i.e., total FOG duration divided by time taken to complete Ziegler’s FOG score). Herman and colleagues found time taken to be an independent predictor of self-reported FOG severity (i.e., total New FOG Questionnaire score) in 71 people with Parkinson’s disease and moderate-to-severe FOG (“On” state: R = 0.26, p = 0.041; “Off” state: R = 0.53, p < 0.001), but this appeared to explain only 6.8% and 28.1% of the variance in the “on” and “off” states respectively.^32^ There was also no significant correlation between time taken and the New FOG Questionnaire’s Part II, which rates FOG severity based on its frequency and duration.^32,35^ Furthermore, 32% of participants (n = 23) were unable to complete the tasks in Ziegler’s FOG score in either the “on” state (i.e., when FOG is typically least severe), “off” state (i.e., when FOG is typically most severe), or both “on” and “off” states, rendering time taken impossible to record in these instances.^32^

The tasks performed in Ziegler’s FOG score may have limited the validity of its timed measure. Content validity, which encompasses concepts such as relevance and comprehensiveness, is uncertain for Ziegler’s FOG score.^36^ Ziegler’s FOG score includes dual-tasking by walking while carrying a tray.^19^ This limits participation to people who do not need a walking aid for independent ambulation. Yet, walking aid use is not uncommon among people with Parkinson’s disease.^7,37^ Ziegler’s FOG score also has a sit-to-stand component which would exclude those who are unable to stand up from a chair without physical assistance.^19^ However, rather than FOG, sit-to-stand ability depends on strength, overall bradykinesia, posture, balance, and cognition.^38^ In contrast to Ziegler’s FOG score, the FOG Severity Tool was deliberately developed through consensus opinion of healthcare professionals for relevance and comprehensiveness.^20^ It allows walking aid use, does not involve standing up from a chair, and may be more successful in triggering FOG given the larger turning angle and smaller width of the narrow-space.^14,20–22,39,40^ Compared to the commonly-used Timed Up and Go,^34^ the FOG Severity Tool had 6.2 times the odds of eliciting FOG.^21^

With the aim of providing a quick and easy-to-use clinical measure, this study investigated if time taken to complete the FOG Severity Tool (or its components) could reflect FOG severity. To further support score interpretation and implementation in routine clinical practice, MIC and time criterion for test discontinuation were calculated for the selected timed measure (i.e., Time To Navigate, TTN).

## Methods

### Participants

Consecutive community-dwelling people with Parkinson’s disease were recruited from outpatient clinics of a tertiary hospital between August 2021 and August 2022. Selection criteria were the age of at least 30 years, ability to ambulate 8-metres without physical assistance (regardless of walking aid use), ability to follow instructions for study procedures, and being without other conditions that severely affected gait. This study received ethics approval (CIRB 2019/2650, HRE2020-0094) and all participants provided written informed consent.

### Study procedures

Demographic information, such as age and disease duration, were collected. Thereafter, participants were assessed with the FOG Severity Tool in a test-retest fashion, with approximately 30-to 60-minutes between testing occasions.^22^ The FOG Severity Tool’s assessment course included the following, performed in both single-task and cognitive dual-task (i.e., simultaneous performance of a rule-reversal task with response of either “big” or “small” to the given numbers which ranged between 1 and 10)^21^ conditions:

1. Six-metre forward-walk at a comfortable pace;
2. On-the-spot turns at a fast pace, twice clockwise and anti-clockwise, performed separately to provide for an opportunity to rest in between (and, consequently, timed separately); and
3. Forward-walk at a comfortable pace through a 50-centimetre narrow space (i.e., just enough to accommodate the width of typical walking frames used locally) created between a high-back chair and a wall.^21^

The full assessment course was video-recorded for post-hoc analysis. Between testing occasions of the FOG Severity Tool, the FOG Questionnaire,^41^ Parkinson Anxiety Scale,^42^ Montreal Cognitive Assessment,^43^ and Movement Disorder Society’s revised Unified Parkinson’s Disease Rating Scale (MDS-UPDRS) Parts II and III were administered.^44^ For perceived change in FOG severity between testing occasions, participants were asked to provide a rating on a 7-point global rating of change scale before commencing the second occasion of the FOG Severity Tool.^45^

### Outcomes

The timed measures were not collected during the actual performance of the FOG Severity Tool as the assessor’s focus was on scoring the tool. Instead, the time taken for each component of the FOG Severity Tool was recorded with a stopwatch during video-analysis. The following were derived from this:

1. Time taken to complete the FOG Severity Tool (i.e., FOG Severity Tool Time) – by summing the time taken for all components of the FOG Severity Tool;
2. Time taken to complete the on-the-spot turning and narrow-space components (i.e., TTN) – by summing the time taken for these components of the FOG Severity Tool in both single-task and dual-task conditions (see Figure 1);
3. Adjusted TTN – as a means of accounting for other impairments that could affect gait timing (e.g., bradykinesia, hypokinesia etc.), by subtracting the time required to complete the single-task six-metre forward-walk from the TTN.

**Figure 1.**
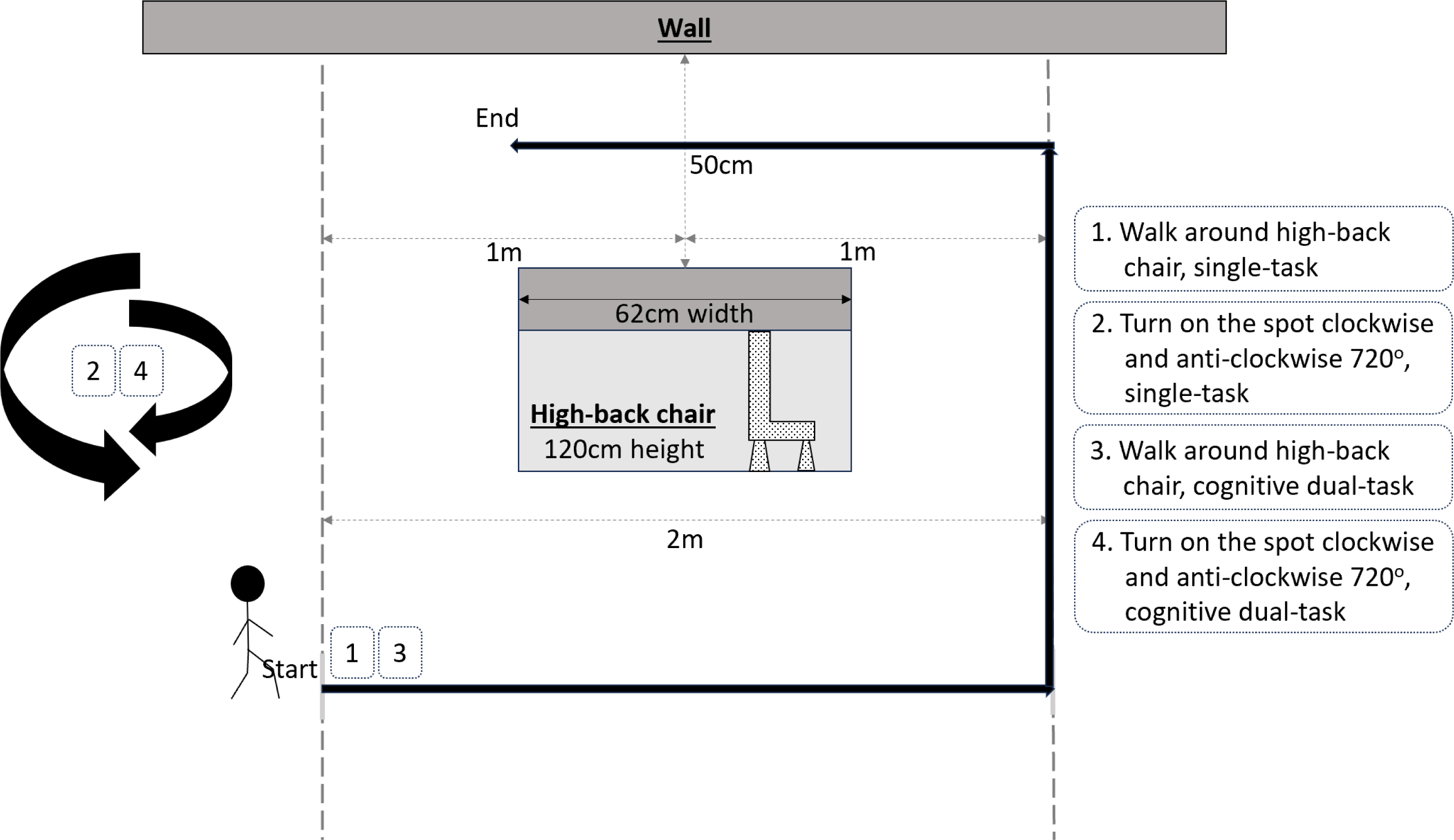
Illustration of the course used for TTN.

### Statistical analysis

For continuous variables, means with standard deviations (SD) and medians with interquartile ranges (IQR) were calculated. For categorical variables, frequencies with percentages were computed. To investigate convergent validity of each timed measure, Spearman’s correlation coefficients with bootstrapped 95% confidence intervals were calculated with three existing FOG severity outcome measures:

1. FOG Questionnaire, which was found to be the best available FOG severity outcome measure based on the COSMIN;^36^
2. Percentage of time spent with FOG (during performance of the complete FOG Severity Tool), which is commonly considered the “gold standard” FOG severity outcome in research;^10^ and
3. The recently-validated FOG Severity Tool–Revised score.^22^

To explore potential time criterion for test discontinuation for the selected timed measure (i.e., TTN), TTN was plotted against the FOG Questionnaire, percentage of time spent with FOG, and the FOG Severity Tool–Revised score using the LOESS (locally estimated scatterplot smoothing) method.^46^

To facilitate the clinical interpretability of the TTN scores, its MIC was calculated with the predictive modelling method developed by Terluin and colleagues,^47^ using the MIC R package.^48^ The 7-point global rating of change scale was the anchor measure, and participants were classified as being importantly improved when they responded “slightly better”, “moderately better”, or “very much better”. The predictive modelling method involved regressing the dichotomised anchor responses on the change in TTN scores and estimating the change in TTN score (i.e., the MIC) that corresponded to a likelihood ratio of 1.0 between the pre- and post-test odds of an “improved” anchor response.^47,49^ To compute 95% CIs for the MIC, percentile bootstrapping (1000 resamples) was used. Notably, as the proportion of improved participants on the global rating of change scale was close to 0.50 (0.48), an adjusted MIC was not calculated.^47^ All analyses were done in R, version 4.2.2 (R Foundation, Vienna, Austria). Statistical significance was set at P < 0.05.

## Results

Forty-one people with Parkinson’s disease were enrolled. Of these, four were unable to complete the FOG Severity Tool on either the first or second testing occasion (9.8%), one declined to stay for a second testing occasion due to a conflicting hospital appointment (2.4%), and one withdrew consent (2.4%). Consequently, data from 35 participants were analysed in this study. Table 1 summarises demographic information and descriptive statistics. Among the 35 participants, 11.4% (n = 4) who had self-reported FOG did not show any FOG during performance of the complete FOG Severity Tool.

**Table 1.** Participants (N = 35) demographics and descriptive results.

| Characteristics | Mean (SD) (25<sup>th</sup>, 50<sup>th</sup>, 75<sup>th</sup> Percentile)<sup>a</sup> |  |
| --- | --- | --- |
| Age, years | 69.7 (9.3) | (64.0, 73.0, 75.0) |
| Sex (male), n (%) | 29 | 82.9 (66.4 – 93.4) % |
| Marital status, n (%) – |  |  |
| Married | 26 | 74.3 (62.9 – 88.8) % |
| Single | 4 | 11.4 (0.0 – 25.9) % |
| Divorced | 3 | 8.6 (0.0 – 23.1) % |
| Widowed | 2 | 5.7 (0.0 – 20.2) % |
| Occupation, n (%) – |  |  |
| Retired | 25 | 71.4 (60.0 – 87.6) % |
| Working | 8 | 22.9 (11.4 – 39.1) % |
| Home-maker | 1 | 2.9 (0.0 – 19.1) % |
| Job-seeking | 1 | 2.9 (0.0 – 19.1) % |
| Lives with, n (%) – |  |  |
| Family | 30 | 85.7 (69.7 – 95.2) % |
| Alone | 5 | 14.3 (4.8 – 30.3) % |
| Mobility status, n (%) – |  |  |
| Community ambulant | 30 | 85.7 (69.7 – 95.2) % |
| Home-bound | 5 | 14.3 (4.8 – 30.3) % |
| Hoehn & Yahr Stage, n (%) – |  |  |
| I | 3 | 8.6 (0.0 – 27.5) % |
| II | 13 | 37.1 (22.9 – 56.1) % |
| III | 15 | 42.9 (28.6 – 61.8) % |
| IV | 4 | 11.4 (0.0 – 30.3) % |
| Medication state during testing, n (%) – |  |  |
| On | 30 | 85.7 (69.7 – 95.2) % |
| Off | 5 | 14.3 (4.8 – 30.3) % |
| Disease duration, years | 4.4 (4.1) | (1.5, 4.0, 6.0) |
| Number of falls in the past 12 months | 3.4 (6.0) | (0.0, 0.0, 2.0) |
| Parkinson Anxiety Scale, total | 9.5 (9.6) | (2.0, 8.0, 16.0) |
| Montreal Cognitive Assessment, total | 25.0 (3.9) | (23.0, 25.0, 27.0) |
| FOG Questionnaire, total | 8.0 (6.4) | (3.0, 8.0, 11.5) |
| MDS-UPDRS II, total | 12.1 (8.2) | (6.0, 12.0, 16.0) |
| MDS-UPDRS III, total | 51.3 (20.3) | (37.5, 53.0, 62.0) |
| FOG Severity Tool-Revised, total | 13.7 (12.2) | (6.0, 9.0, 17.5) |
| FOG Severity Tool Time, seconds | 139.5 (117.7) | (66.1, 85.8, 145.6) |
| TTN, seconds | 119.2 (105.4) | (54.8, 74.8, 129.9) |
| Adjusted TTN, seconds | 110.2 (100.4) | (49.5, 69.9, 122.9) |

Abbreviations: FOG – Freezing of gait; MDS-UPDRS II – Movement Disorder Society's revised Unified Parkinson's Disease Rating Scale Part II; MDS-UPDRS III – Movement Disorder Society's revised Unified Parkinson's Disease Rating Scale Part III
*Note.* <sup>a</sup>Values are mean (SD) (25<sup>th</sup>, 50<sup>th</sup>, 75<sup>th</sup> percentile) unless stated otherwise. Variables can be interpreted as being normally distributed if the median and mean are similar and the 25<sup>th</sup> and 75<sup>th</sup> percentiles are symmetrical compared to the median. For percentages, 95% confidence intervals are presented in brackets.

FOG Severity Tool Time, TTN, and adjusted TTN each correlated moderately with both the FOG Questionnaire (Spearman’s rho = 0.61 – 0.63) and the percentage of time spent with FOG during performance of the complete FOG Severity Tool (Spearman’s rho = 0.67 – 0.68) (see Table 2). Correlations between the timed measures and the FOG Severity Tool–Revised score were very high (Spearman’s rho = 0.93 – 0.94) (see Table 2). Given the similar performance of the three timed measures, the one which required the fewest number of assessment components and computation steps was selected – TTN.

**Table 2.** Correlations with Freezing of Gait Questionnaire and percentage of time spent with freezing of gait.

| <u>Freezing of Gait (FOG) Questionnaire</u> | <u>Spearman's rho (95% confidence interval)</u> |
| --- | --- |
| TTN | 0.63 (0.37 – 0.81) |
| Adjusted TTN<sup>a</sup> | 0.63 (0.37 – 0.80) |
| FOG Severity Tool Time | 0.61 (0.33 – 0.79) |
| Percentage of time spent with FOG<sup>b</sup> | 0.57 (0.28 – 0.78) |
| <u>Percentage of time spent with FOG</u> |  |
| TTN | 0.68 (0.43 – 0.84) |
| Adjusted TTN | 0.68 (0.46 – 0.84) |
| FOG Severity Tool Time | 0.67 (0.44 – 0.82) |
| FOG Severity Tool-Revised score | 0.67 (0.44 – 0.82) |
| <u>FOG Severity Tool-Revised score</u> |  |
| TTN | 0.94 (0.85 – 0.97) |
| Adjusted TTN | 0.94 (0.84 – 0.97) |
| FOG Severity Tool Time | 0.93 (0.84 – 0.97) |

Abbreviations: FOG – Freezing of gait
*Note.* <sup>a</sup>Time taken to walk 6-metres subtracted from TTN; <sup>b</sup>Percentage of time spent with FOG in the FOG Severity Tool's assessment course.

LOESS plots of the TTN against the FOG Questionnaire, percentage of time spent with FOG during performance of the complete FOG Severity Tool, and FOG Severity Tool–Revised score are presented in Figure 2 (i.e., Figures 2a, 2b, and 2c respectively). TTN was nonlinearly related to FOG severity such that a positive relationship was observed in the first 300 seconds (i.e., five minutes), beyond which the association plateaued, with additional time beyond 300 seconds no longer producing appreciable changes in FOG severity scores. This suggested that the TTN test could be discontinued after 300 seconds. MIC for the TTN was a time reduction of 15.8 seconds (95% CI 3.2 to 28.7). This translated to 13.3% and 21.1% of the mean and median TTN respectively.

**Figure 2.**
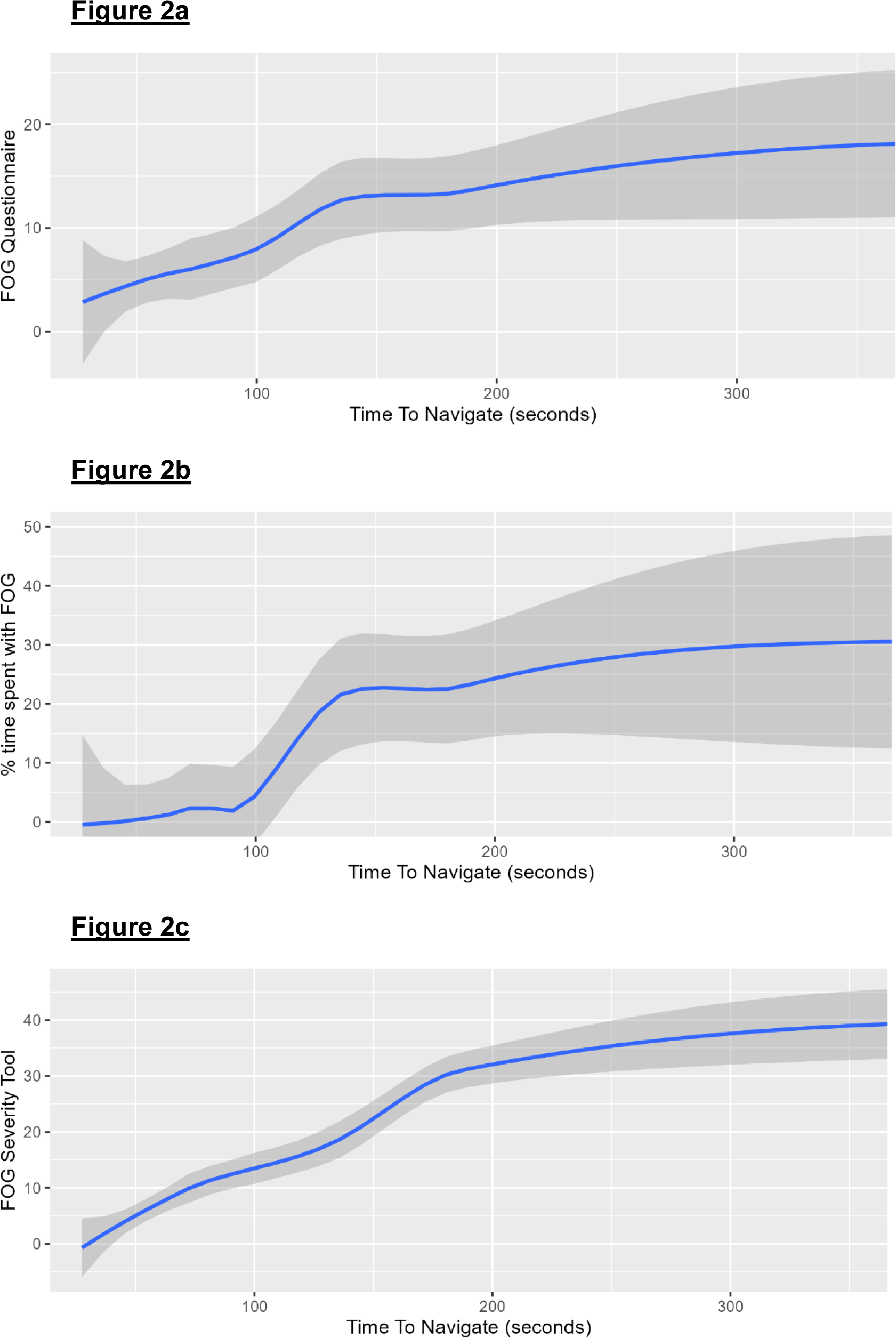
LOESS smoothing curves (with 95% CI) illustrating the nonlinear associations of TTN with the (a) Freezing of gait (FOG) Questionnaire, (b) percentage of time spent with FOG during performance of the complete FOG Severity Tool, and (c) FOG Severity Tool–Revised score.

## Discussion

This study evaluated timed measures based on the FOG Severity Tool and found the TTN may be considered for a quick and easy-to-use clinical measure. The TTN is a valid, feasible, and interpretable test of FOG severity. Similar to the FOG Severity Tool Time and adjusted TTN, TTN correlated moderately with both the FOG Questionnaire and percentage of time spent with FOG during performance of the complete FOG Severity Tool. With the FOG Severity Tool–Revised score, which previously demonstrated sufficient criterion-related validity against the FOG Questionnaire,^22^ correlation was very high. To further support implementation in routine clinical practice, a time criterion for test discontinuation (i.e., 300 seconds) was suggested to facilitate time efficiency and feasibility whilst an estimate for MIC (i.e., a reduction in timing by 15 seconds) was calculated to facilitate clinical interpretability – both of which are regarded by the COSMIN as important factors for outcome measure selection.^23,24^

The challenge of translation from research to routine clinical practice is known, but there has been no easy route identified for achieving this.9 It has been suggested that focus groups should be conducted with clinicians to determine from their perspective what information is important for FOG severity.9 We developed the FOG Severity Tool through a Delphi study, based on the consensus opinion of 28 medical specialists, advanced practice nurses, and specialised physiotherapists.20 Apart from informing the items for inclusion in the FOG Severity Tool, the participating healthcare professionals also gave insight into factors considered important by clinicians for uptake – practicality, which consisted of efficiency and ease of use; and usefulness, which included level of detail and comparability to FOG experience in the home setting.20

In terms of practicality, the TTN was more efficient than the FOG Severity Tool Time, the adjusted TTN, and even the FOG Severity Tool–Revised score – it had fewer assessment components, required fewer computational steps, had median completion time of 74.8 seconds, and could be stopped at about 300 seconds (i.e., 5 minutes). In contrast, the FOG Severity Tool took an average of 6.5 minutes to complete and Ziegler’s FOG score needed up to 15 minutes.^19,21^ There has been no recommended cut-off time for Ziegler’s FOG score though previous studies have recorded participants taking over 100 seconds for the third condition alone.^32^ *(A summary of completion times and interpretation thresholds for objective FOG severity outcome measures is provided in Supplementary Material 1.)* Without a need to familiarise with a clinical rating scale, the TTN may also be easier to use. Like the Timed Up and Go compared to the original Get Up and Go test,^50^ considerably less expertise and training is needed to administer the TTN test.^32,34^

With its very high correlation, the TTN could be an attractive alternative to the FOG Severity Tool–Revised.^22^ For comprehensiveness and relevance, the items within the FOG Severity Tool were informed by experienced healthcare professionals and were specifically intended to differentiate between levels of FOG severity.^20,21^ Though the TTN represents fewer assessment components, correlation strength was comparable to that of timing all assessment components (i.e., FOG Severity Tool Time). However, retaining only the assessment components more likely to trigger FOG in the TTN may mean people with more severe FOG have no measured outcome, since an inability to complete the assessment components means no time taken can be recorded. This affected about 10% (n = 4) of the enrolled participants in this study. While the advantage of the FOG Severity Tool–Revised (or any equivalent clinician-rated scale) is that such participants still receive a score of 9 for each assessment component they are unable to complete, in the clinical setting the missing TTN score may be less important since improvement would still be defined based on the recovered ability to complete the assessment components. Moreover, in the research setting, a potential approach to handle the missing data of participants who are unable to complete the TTN test may be to censor the TTN values at 300 seconds.

Another consideration for usefulness was comparability to FOG experience in the home setting.^20^ It has been suggested that objective assessments for FOG severity should ideally be carried out in the natural environment (e.g., the home setting) to prevent shifts from automatic gait control toward attention, which reduces FOG.^9^ Yet, there are currently no standardised assessment protocols for home-based evaluations of FOG severity. Although FOG can be monitored in daily-life with wearable sensors, a standardised assessment course may be necessary for specific pre- and post-intervention comparisons since there is day-to-day variability.^29^ The fewer assessment components and minimal equipment used in the TTN may help execution within the home environment. Further studies could explore validity of timing within the home with use of alternative equipment (e.g., a coffee table in place of the high-back chair).

### Limitations

This study had limitations. Firstly, while using a timed measure like the TTN to reflect FOG severity may offer efficiency and ease of use, it entailed a trade-off since changes in time taken may not be attributed to FOG specifically.^33^ Changes in cognition over time may influence TTN, especially its dual-task assessment components.^51^ Changes in cognitive assessment scores may need to be considered for interpretation of longitudinal changes in TTN. Secondly, medication state could not be controlled for because the research began when COVID-19 restrictions were still in place, resulting in an inability to dictate appointment times and a limit placed on appointment durations. The specific effects of medication state on the TTN may need further investigation. In spite of the majority of study participants being in the “on” state, correlation between the TTN and FOG Questionnaire was moderate. This is in contrast to the low correlation between the time taken to complete Ziegler’s FOG score in the “on” state and the New FOG Questionnaire.^32^ Thirdly, the timed measures were obtained using a stopwatch during video-analysis – whether assessors can record the TTN time with a stopwatch during the actual TTN test performance, without needing a video-recording, remains to be demonstrated. Finally, the participants in this study had milder FOG severity. Validity, cut-off time, and MIC of the TTN in people with more severe FOG remains to be investigated. In addition, though the TTN showed promise in a sample comparable in size to published validation studies for other FOG severity outcome measures,^16,19^ evaluation in a larger sample is still needed for firm conclusions to be drawn.

## Conclusion

This study aimed to provide a quick and easy-to-use clinical measure for reflecting FOG severity – the TTN. In this initial sample of people with Parkinson’s disease, the TTN demonstrated moderate correlations with the FOG Questionnaire and percentage of time spent with FOG during performance of the complete FOG Severity Tool. Given very high correlations with the recently-validated FOG Severity Tool–Revised score, the TTN may be a convenient alternative. For time efficiency, the TTN may be stopped at 300 seconds (i.e., 5 minutes), with a decrease in TTN of 15.4 seconds considered minimal improvement in FOG severity.

## Data Availability

All data produced in the present study are available upon reasonable request to the authors.

## Abbreviations

FOG: Freezing of gait
MIC: Minimal important change
TTN: Time To Navigate

## Ethics approval

This study was approved by the ethics committees of the relevant institutions (CIRB 2019/2650, HRE2020-0094).

