## Supplementary Material 1 for "Time To Navigate (TTN): A practical objective clinical measure for freezing of gait severity in people with Parkinson’s disease"

**Supplementary Material 1.** Completion times and interpretation thresholds, where available, for objective freezing of gait severity outcome measures with reported validity and reliability

| Outcome measure | Completion Time | Interpretation Thresholds |
|---|---|---|
| | *Clinician-rated outcome measures* | |
| Freezing of Gait Severity Tool[21] | Median (IQR): 6.5 (4.5) min<br>Range: 3.5 to 16.0 min | Unknown |
| Cremers and colleagues' Dynamic Parkinson Gait Scale[16] | "Average" 4 to 8 min | Unknown |
| Gavriliuc and colleagues' four-point scale[17] | Unknown | Unknown |
| Kim and colleagues' four-point scale[18] | Unknown | Unknown |
| Ziegler and colleagues' Freezing of Gait Score[19] | Uncertain – estimated up to 15 min | Minimal important change *(ROC method)*:<br>- Improvement = 3<br>*(Sensitivity: 0.67, Specificity: 0.96)*<br>- Worsening = 5<br>*(Sensitivity 0.47, Specificity: 0.96)*[a] |
| | *Direct measures to reflect freezing of gait severity* | |
| Time To Navigate | Median (IQR): 74.8 (75.1) s *or 1.2 (1.3) min* | Minimal important change:<br>15.8 seconds reduction in timing<br>*(95% CI 3.2 to 28.7)* |
| Time to complete Ziegler and colleagues' Freezing of Gait Score[32] | "On" medication state –<br>Condition 1:<br>Median (IQR): 38.0 (27.2) s<br>Condition 2:<br>Median (IQR): 38.9 (27.1) s<br>Condition 3:<br>Median (IQR): 56.3 (38.8) s<br><br>"Off" medication state –<br>Condition 1:<br>Median (IQR): 45.6 (32.0) s<br>Condition 2:<br>Median (IQR): 49.0 (33.7) s<br>Condition 3:<br>Median (IQR): 71.0 (56.3) s<br><br>"On" medication state:<br>133.2 s *or 2.2 min*<br><br>"Off" medication state:<br>165.6 s *or 2.8 min* | Unknown |
| Percentage of time spent with freezing of gait in Timed Up and Go[26] | Uncertain – estimated at under 3.2 min for each video-analysis alone | Unknown |

*Note.* Abbreviations – ROC: Receiver-operating characteristics curve

[a]Reference: Fietzek UM, Schulz SJ, Ziegler K, Ceballos-Baumann AO. The Minimal Clinically Relevant Change of the FOG Score. J Parkinsons Dis. 2020;10(1):325-332. doi:10.3233/JPD-191783.
